# Removal of Free Liquid Layer from Liquid-Infused Catheters Reduces Silicone Loss into the Environment while Maintaining Adhesion Resistance

**DOI:** 10.1101/2023.09.14.23295548

**Authors:** Chun Ki Fong, Marissa Jeme Andersen, Emma Kunesh, Evan Leonard, Donovan Durand, Rachel Coombs, Ana Lidia Flores-Mireles, Caitlin Howell

## Abstract

Silicone urinary catheters infused with silicone liquid offer an effective alternative to antibiotic coatings, reducing microbial adhesion while decreasing bladder colonization and systemic dissemination. However, loss of free silicone liquid from the surface into the host system is undesirable. To reduce the potential for liquid loss, free silicone liquid was removed from the surface of liquid-infused catheters by either removing excess liquid from fully infused samples or by partial infusion. The effect on bacterial and host protein adhesion was then assessed. Removing the free liquid from fully infused samples resulted in a ∼64% decrease in liquid loss into the environment compared to controls, with no significant increase in deposition of the host protein fibrinogen or the adhesion of the common uropathogen *Enterococcus faecalis*. Partially infusing samples decreased liquid loss as total liquid content decreased, with samples infused to 70-80% of their maximum capacity showing a ∼85% reduction in liquid loss compared to fully infused controls. Furthermore, samples above 70% infusion showed no significant increase in fibrinogen or *E. faecalis* adhesion. Together, the results suggest that eliminating free liquid layer, mechanically or through partial infusion, can reduce liquid loss from liquid-infused catheters while preserving functionality.

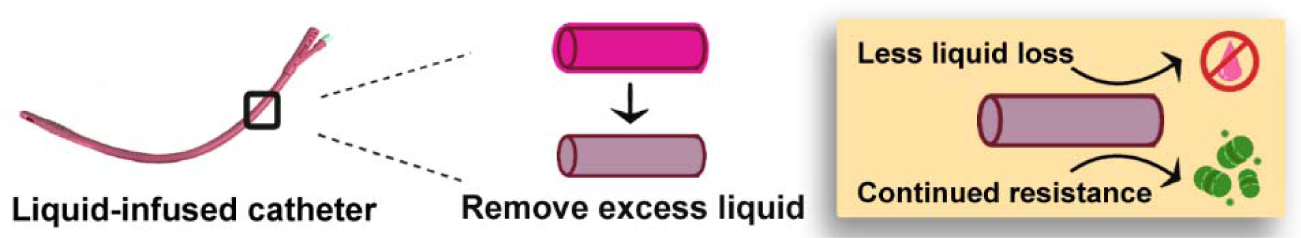

## 1. Introduction

Catheter-associated urinary tract infection (CAUTI) is one of the most common types of infection developed in hospitals, with over 400,000 cases reported annually in the United States alone. ^[1-3]^ Catheterized patients have higher mortality rates within a year compared to catheter-free patients with similar medical histories.^[4]^ This increased mortality can be attributed to complications resulting from pathogen colonization on catheter material, including well-known CAUTI-causing agents such as *Escherichia coli* and *Enterococcus faecalis*.^[4]^ In addition, reports of antimicrobial-resistant strains of both bacteria and fungi associated with CAUTI are increasing each year, worsening the financial burden on and life quality of patients.^[5-6]^ Therefore, new, robust, and easily deployable solutions that do not rely on antimicrobial compounds are needed.

One promising approach to reducing the initiation and spread of CAUTI without antibiotics has recently been demonstrated *in vivo*: the application of silicone liquid via liquid infusion to commercially available all-silicone catheter materials. In solid silicone polymers, liquid infusion is accomplished when molecules of a compatible liquid, such as silicone liquid, interpenetrate in between the crosslinked chains of the solid.^[7-10]^ Full infusion is generally understood to create a layer of free silicone liquid on the catheter surface, imparting antifouling properties that prevent the irreversible adherence of contaminants.^[9; 11-16]^ Silicone liquid-infused polymers have shown promising results in both *in vitro* and *in vivo* settings, effectively reducing adhesion by a range of medically relevant proteins and microorganisms.^[7;12-13;17-20]^ In particular, a recent *in vivo* study by Andersen *et al* has shown that these materials can reduce the adhesion of fibrinogen, a host protein which facilitates the attachment and proliferation of microorganisms, in a murine model of CAUTI. ^[13]^ This work demonstrated that preventing the adhesion of fibrinogen decreased subsequent attachment by six of the most prevalent pathogens in CAUTI, as well as significantly reducing bladder colonization and their dissemination to other organs. These results point to the importance of deposited proteins in infection and the role that liquid-infused polymers can play as a robust and easily applicable method for reducing protein deposition on surfaces.

While the *in vivo* reduction of protein adhesion by liquid-infused urinary catheters as a mechanism to control infection is promising, there is concern over the potential for loss of the free liquid into the host system. ^[21-23]^ Multiple studies focused on ocular tamponades and connective tissue have demonstrated the potential for adverse effects of silicone liquid leakage into the host tissue including heightened inflammatory cell response and antibody production.^[1-4,9,24]^ It is currently believed that the immune response is primarily due to the formation of protein aggregates around silicone liquid droplets. *In vivo* studies have shown elevated concentrations of antidrug antibodies in plasma in the presence of silicone liquid-protein complexes.^[22-23]^ Recent studies have also shown that the presence of liquid silicone droplets can trigger the formation of adherent cell masses similar to what is observed in granuloma formation.^[25]^ However, the amount of free silicone liquid necessary to induce an immune response in any given host is not well understood and likely varies from individual to individual. It is estimated that the volume of silicone liquid that coats clinically used prefilled siliconized syringes can reach between 0.6–1 mg/m,^[26]^ with 0.11± 0.03 mg/ml of that volume generally released into the blood stream,^[23]^ suggesting that a limited exposure to free silicone liquid in the host system is tolerable.

Previous studies have demonstrated that repeated exposure to an air/water interface is an effective method for removing the free liquid layer from infused systems, as the lower-surface-energy liquid will spontaneously move to cover the higher-surface-energy water interface and be carried away into the environment.^[8,27-28]^ In urinary catheters, the intermittent flow of urine over the surface of the catheter is very likely to result in the loss of free silicone liquid from liquid-infused surfaces. At the same time, repeated contact of the exterior surface of the catheter with urethral and bladder tissue is likely to physically remove free liquid present there.^[29]^ It is therefore critical to understand the potential for surface liquid loss in liquid-infused catheters and explore methods of reducing such loss while continuing to preserve their antifouling capabilities.

The goal of this work was to explore the effect of the removal of the free liquid layer from the surface of infused catheter samples on liquid loss into the environment. We further aimed to measure how such a treatment would affect the adhesion of host protein and bacteria relevant to CAUTI. Here we show that surface liquid removal significantly decreases silicone liquid lost into the environment without disrupting protein- and bacterial-resistance as long as a certain volume of silicone liquid (∼80%) remains in the system. Our results will help to further the use of liquid-infused urinary catheters and other devices as safe and effective alternatives to antibiotic coatings in medical settings.

## 2. Results

### 2.1. Removal of the free liquid layer does not significantly affect surface slipperiness

To better understand the role of the free liquid layer in a protein and bacterial adhesion system, we first fabricated infused catheter samples by immersing them in 20 centistoke (cSt) trimethoxy-terminated silicone liquid until complete saturation was achieved, indicated by a plateau in weight gain.^[8,13]^ Samples were then removed from the liquid and all excess liquid was allowed to drain from the surface, these samples were considered to have an intact free liquid layer (LL). A subset of catheter samples was then subjected to removal of the free liquid layer (ØLL) via absorption of the liquid from both interior and exterior surfaces by light contact with a cellulosic wipe **(Figure 1A)**. Unlike previous reports where the free liquid layer was removed via rinsing with water, ^[16,30-31]^ this treatment was intended to deplete the surface layer to the point where it would not substantially increase via syneresis, defined as free silicone liquid molecules migrating to the surface of the material, over the duration of the experiments (<48h), as has been recently reported to occur in infused silicones. ^[16,32]^

**Figure 1.**
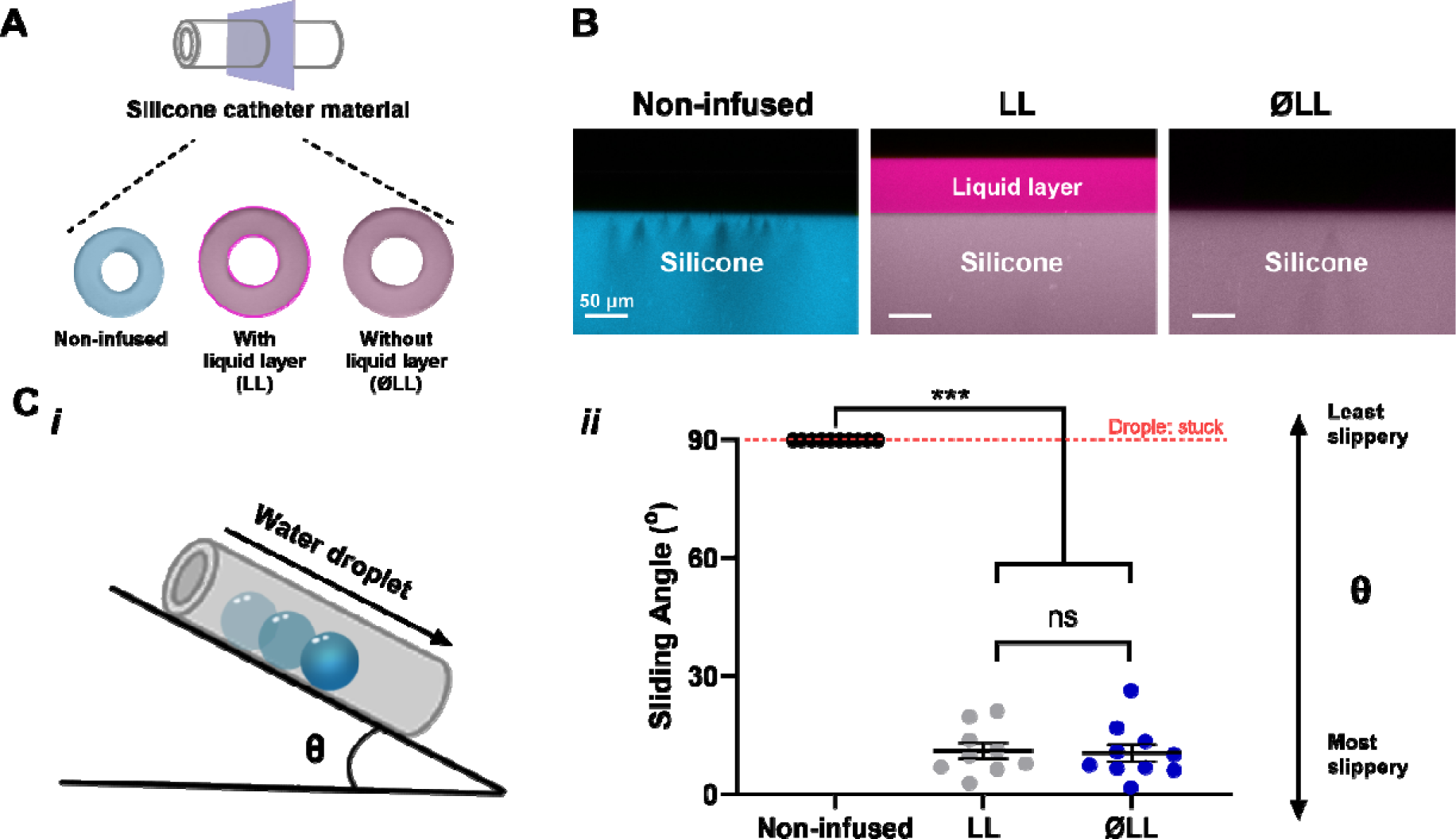
**(A)** Schematic of silicone catheter materials fabricated with or without a liquid layer (LL; ØLL). **(B)** Confocal image analysis of: non-infused silicone catheter material; silicone catheter material with and without liquid layer. Scale bar = 50 µm. **(C*i*)** Schematic of the sliding angle test to characterize slippery surface. **(C*ii*)** Sliding angle test of non-infused silicone catheter material, and silicone catheter material with (LL) or without liquid layer (ØLL). In all graphs presented, the error bars represent the standard error of the mean. Statistical significance between the groups was evaluated using ANOVA test. *** = P < 0.0005 and ns = not significant.

The results of removing the free liquid layer from the surface were visualized using confocal microscopy with different fluorescent dyes for the solid silicone and liquid silicone **(Figure 1B)**. ^[15,33]^ The images of LL samples showed a layer ∼60 µm in thickness, in agreement with previous reports on similar systems.^[15]^ In contrast, ØLL samples showed a marked reduction of the free liquid layer to a value below what could be observed using this technique (≤500nm), confirming the successful removal of nearly all, if not all, the free liquid at the sample surface.

To initially explore the effect of free liquid removal on surface properties, we performed sliding angle tests **(Figure 1C*i*)** where the critical angle at which a water droplet starts to slide down the sample. For liquid-infused surfaces, sliding angle serves as an indicator of the fluid repellency of the surface and therefore the likelihood of resisting adhesion. ^[12,34-37]^ We observed no difference in sliding angle between the LL and ØLL samples **(Figure 1Cii**, P >0.9999). Measurements of the droplet sliding velocity,^[17]^ which provide an indication of surface uniformity, yielded similar results **(Figure S2A)**.

Based on previously published studies, it is generally expected that samples with low sliding angles and high sliding rates would result in the most effective antifouling surfaces. ^[9,15,38]^ With these findings, we conclude that infused silicone catheter materials without a liquid layer exhibits a comparable antifouling surface to the silicone catheter material with a liquid layer.

### 2.2 Removal of free liquid layer significantly reduces liquid loss into the environment

To compare the quantity of liquid that could be lost into the environment in LL vs ØLL samples, catheter sections infused with dyed silicone liquid underwent repeated passage through an air-water interface to strip away the surface liquid layer. The liquid extracted from the infused silicone catheter material into the water was extracted into toluene and subsequently measured for concentration using a spectrophotometer and quantified via a standard curve (**Figure S3**). The results are shown in **Figure 2**. LL samples were found to lose 0.05± 0.01 µL of silicone liquid/mm of sample length, while ØLL samples lost significantly less, 0.02 ± 0.006 µL of liquid/mm (P = 0.038).

**Figure 2.**
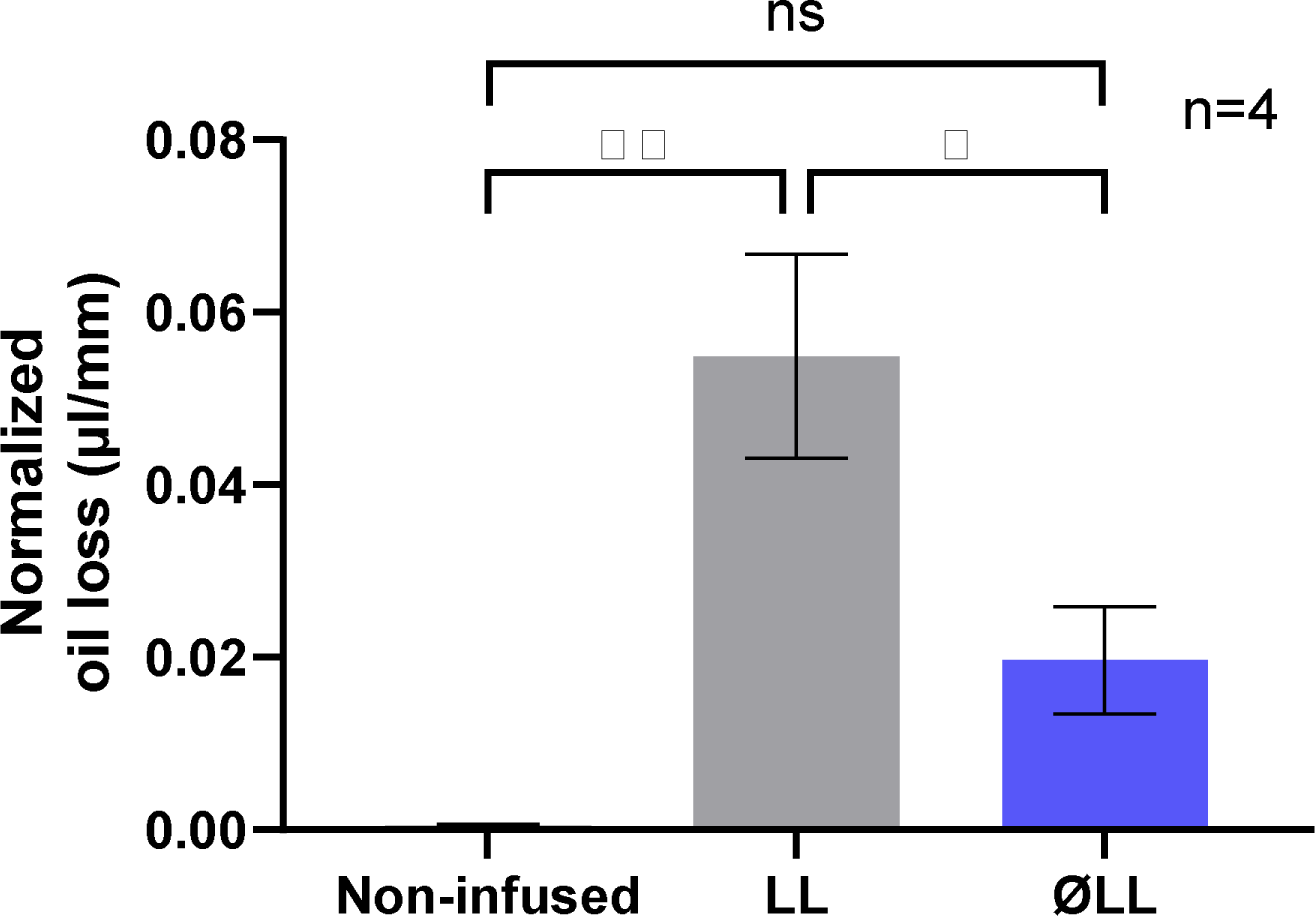
Liquid loss comparison between non-infused silicone catheter samples; silicone catheter samples with and without liquid layer. The error bars represent the standard error of the mean. Statistical significance between the groups was evaluated using ANOVA test. ** = P < 0.01; * = P < 0.05; and ns = not significant.

### 2.3 Exploration of foulant repelling ability of silicone catheter materials without liquid overlayer

Previous investigations of liquid-infused silicone have suggested that a thin, stable free liquid layer is critical to successful fouling resistance, as it acts as a physical barrier and reduces the force required to release attached fouling organisms.^[14,39]^ To assess if removal of the free liquid layer in our system impacted the ability of foulants relevant to CAUTI to adhere to the surface, we incubated both LL and ØLL samples with the host protein fibrinogen and the bacterium *E. faecalis*. The results (**Figure 3)**, showed that both LL and ØLL samples effectively resisted fibrinogen and *E. faecalis* adhesion, showing significantly less surface attachment compared to controls (Fibrinogen: P < 0.0001 and P = 0.0001, respectively; *E. faecalis*: P < 0.0001 and P = 0.009, respectively). Moreover, the results showed no significant difference in either fibrinogen or *E. faecalis* adhesion between LL and ØLL samples (P > 0.9999 and P = 0.25, respectively), suggesting that removing the free liquid overlayer does not have a significant impact on the material’s ability to resist fouling in terms of fibrinogen and pathogen adhesion.

**Figure 3.**
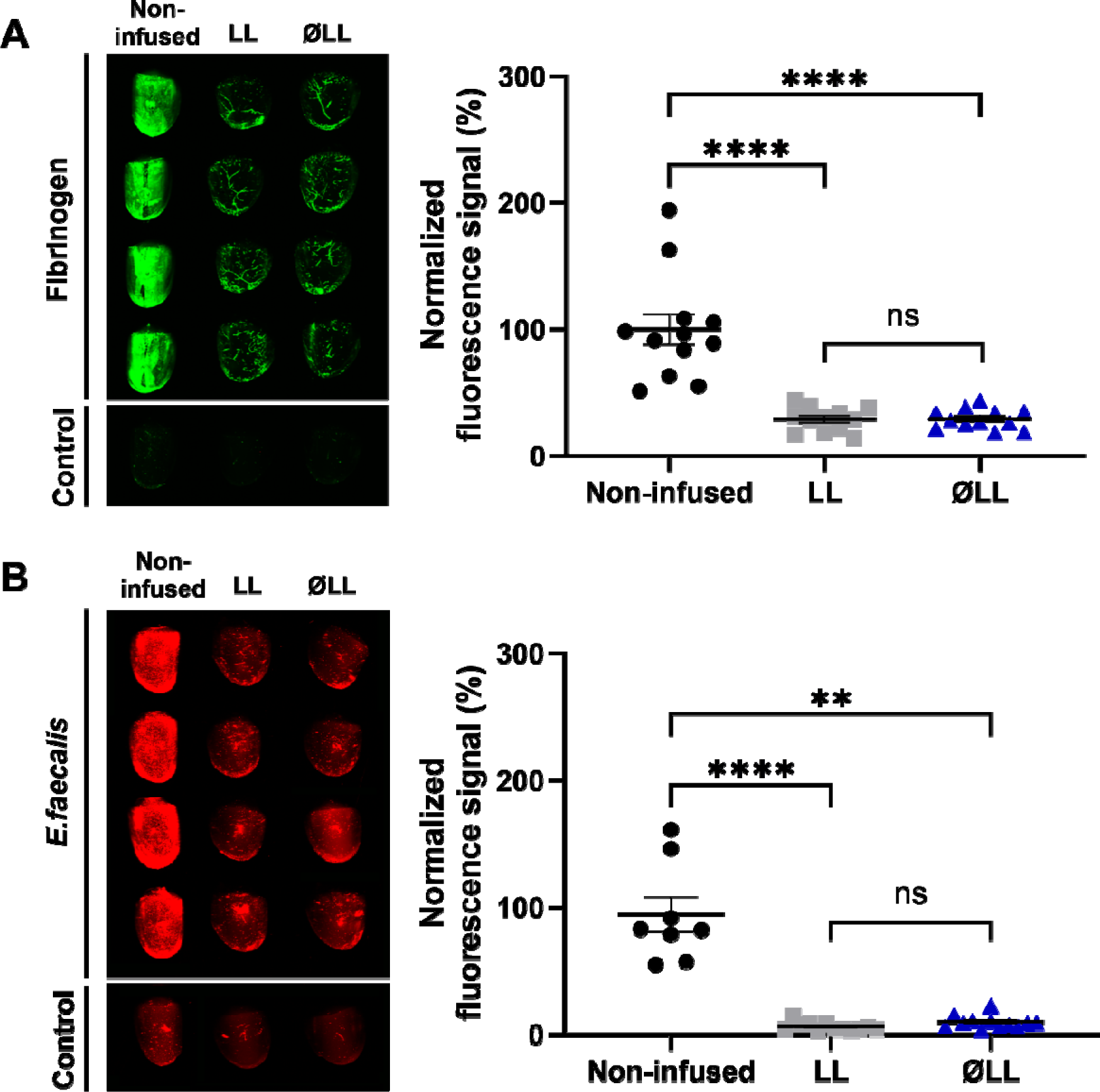
Comparison of **(A)** fibrinogen or **(B)** *E. faecalis* adhesion levels on silicone urinary catheter material that are non-infused, with (LL), or without (ØLL) a liquid layer. The images on the left show the visualization of the samples while the plots on the right show the fluorescence quantification. At least three replicates with n = 4-5 each were conducted. In all graphs presented, the error bars represent the standard error of the mean. Statistical significance between the groups was assessed using ANOVA, with **** = P < 0.0001; ** = P<0.005; and ns = not significant.

### 2.4 Reducing the amount of liquid infused in silicone catheter material

Research has demonstrated that infused silicone polymers can undergo a process called syneresis, in which unbound oligomers or silicone liquid molecules slowly migrate to the surface of the material, re-forming a free liquid layer which is then available to be lost into the environment. ^[16,30,39]^ However, this can be minimized by reducing the amount of free silicone liquid in the system. ^[31]^ We therefore hypothesized that by only partially infusing catheter samples, we could even further reduce the amount of free silicone liquid that could be lost.

Infused materials exhibit a progressive weight increase and swelling during the infusion process until they reach a state of saturation, referred to as full infusion. ^[9]^ The degree of swelling (Q) in the silicone catheter material resulting from the absorption of a solvent such as silicone liquid can be quantitatively described as *Q = w_s_/w_d_*, where *w_s_* represents the weight of the sample after swelling and *w_d_* represents the weight of the dry sample.^[31]^ For ease of comparison, we define the percentage of the maximum degree of swelling (*Q_max_*) as:

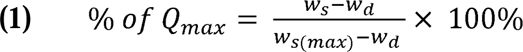

where *w_s_*_(*max*)_ represents the maximum degree of swelling.

In our system, partial infusion was achieved by removing the catheter samples at defined points during the infusion process, as shown in **Figure 4A**, followed by absorbing excess liquid from the surface to prevent further infusion. The result (**Figure 4B**) was a series of well-defined groups of samples with increasingly less free liquid distributed throughout the matrix. Specifically, samples infused for 0.5 hours resulted in 17.4±0.99% of Q_max_, those infused for 6.5 hours achieved 45.25±0.94% of Q_max_, and samples infused for 30.5 hours reached 81.98±1.08% of Q_max_. Samples were considered fully infused if they were removed from the liquid bath at >96h and had a % Q_max_ value of >90.0%.

**Figure 4.**
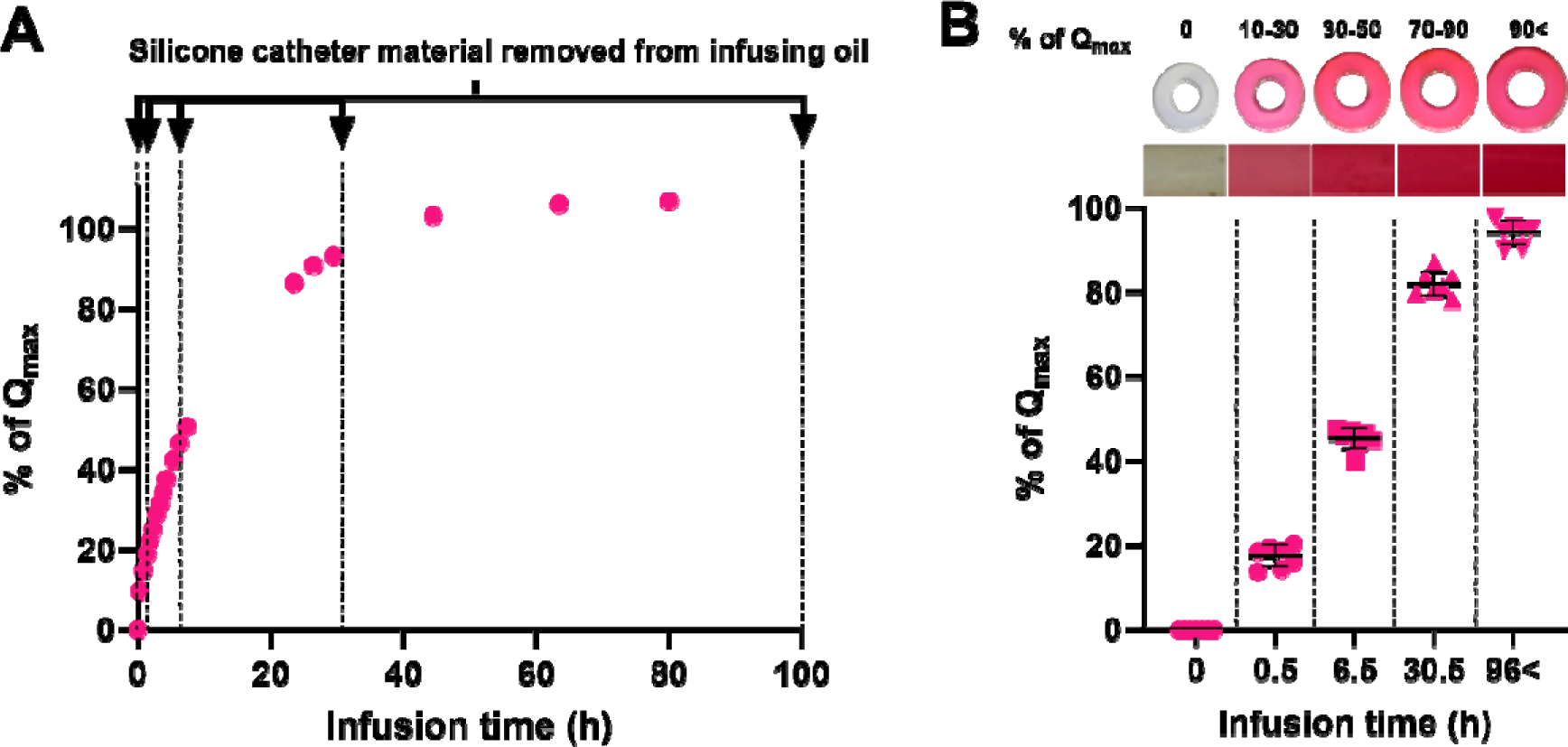
**(A)** Various levels of % of Q_max_ vs infusion time. The arrows denote the timepoint when silicone catheter materials were removed from infusing silicone liquid. **(B)** (Top) Images of sample cross sections in which the infusing liquid was dyed for visualization of the distribution throughout the material. Close-up images of the color are shown in boxes underneath. (Bottom) Silicone catheter materials infused for 0.5; 6.5; 30.5 and >96 hours and their resulting % of Q_max_.

### 2.5 Functionality of partially infused silicone catheter material is comparable to fully infused material and reduces liquid loss

Partially infused samples were tested for both sliding angle and the potential for silicone liquid loss (**Figure 5**). The results of the sliding angle test (**Figure 5A**) showed that as the % of Q_max_ value increases, (*i.e.* a higher amount of free silicone liquid is present in the system), the sliding angle gradually decreases. Samples at 30–50% of Q_max_ already showed a significant difference in sliding angle compared to non-infused controls (P = 0.03). Although samples infused to 70–90% of their Q_max_ showed somewhat higher values than fully infused samples (24.71±2.54 compared to 8.41±1.20, respectively), the difference was found to be non-significant (P > 0.9999). In the droplet velocity test, a similar outcome was observed **(Figure S2)**.

**Figure 5.**
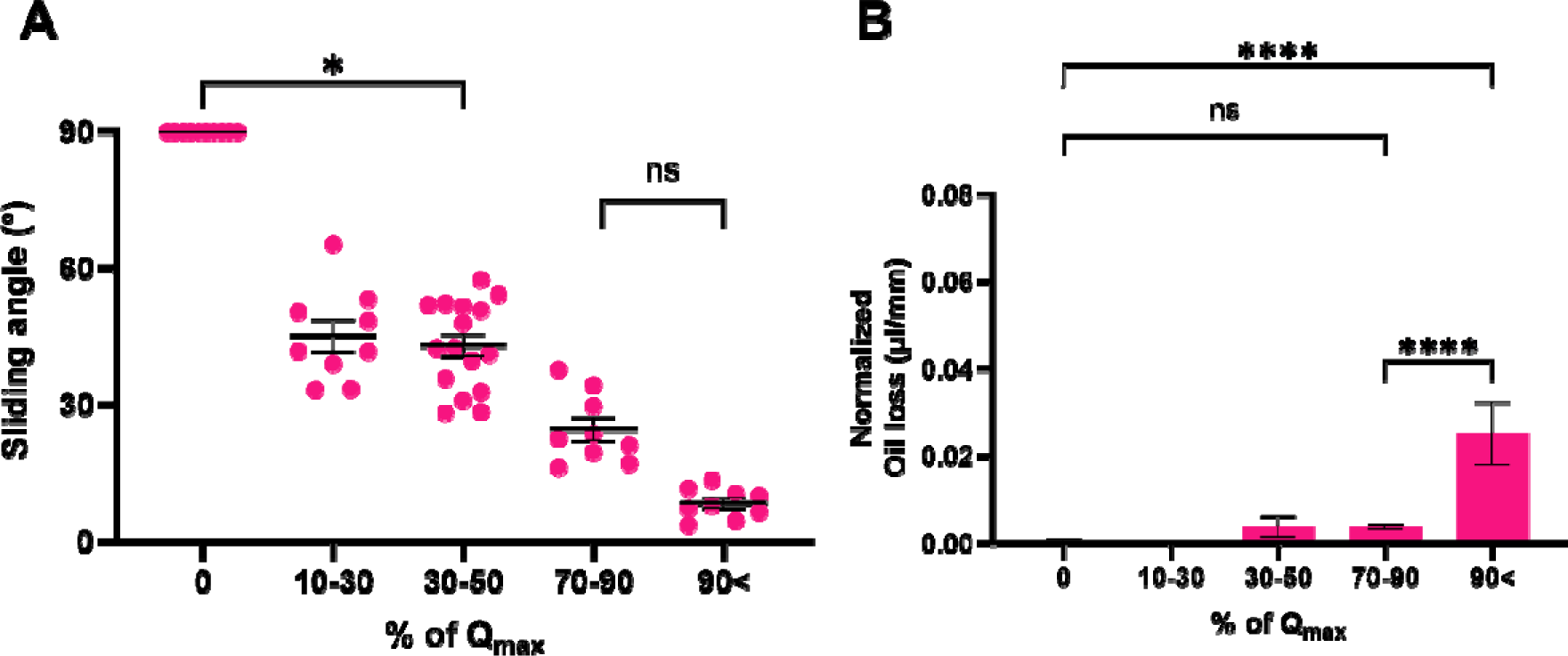
**(A)** Sliding angle of a water droplet on silicone catheter samples with varying % of Q_max_ values. **(B)** Amount of liquid loss per mm of sample’s length for the partially infused silicone catheter samples n=3-7. Statistical significance between the groups was assessed using ANOVA **** = P < 0.0001; * = P < 0.05 and ns = not significant

To investigate the liquid loss levels of partially infused silicone catheter materials, samples were repeatedly exposed to an air-water interface to strip away surface liquid. The results are shown in **Figure 5B**. Fully infused samples were found to lose 0.025±0.004 µl of liquid, while samples at 70–90% of Q_max_ lost significantly less at 0.004±0.0004 µl (P < 0.0001). Samples at 30–50% of Q_max_ lost 0.004 ±0.0013 µl of liquid, which was not significantly different from samples at 70– 90% of Q_max_ (P > 0.9999). Neither samples at 30–50% nor 70–90% of Q_max_ were found to be significantly different from non-infused controls (P = 0.50 and 0.52, respectively), unlike fully infused samples, which were significantly higher (P < 0.0001).

### 2.6 Significant reduction in protein and bacteria adhesion is achievable without full infusion of silicone catheter material

To assess the effectiveness of partially infused silicone catheter material in repelling fouling agents, we again incubated the samples with either fibrinogen or *E. faecalis* followed by immunolabeling to assess the levels of adhesion **(Figure 6)**. The results indicate that, as the infusion level increased, the adhesion levels of both fibrinogen and *E. faecalis* gradually decreased. Notably, the first significant difference was observed between non-infused catheters and those infused to the 70–90% of Q_max_ (P < 0.0001 for both fibrinogen and *E. faecalis*). In addition, silicone catheter material infused to 70–90% of Q_max_ did not show a significant difference compared to fully infused materials (P = 0.19 for fibrinogen; P > 0.9999 for *E. faecalis*).

**Figure 6.**
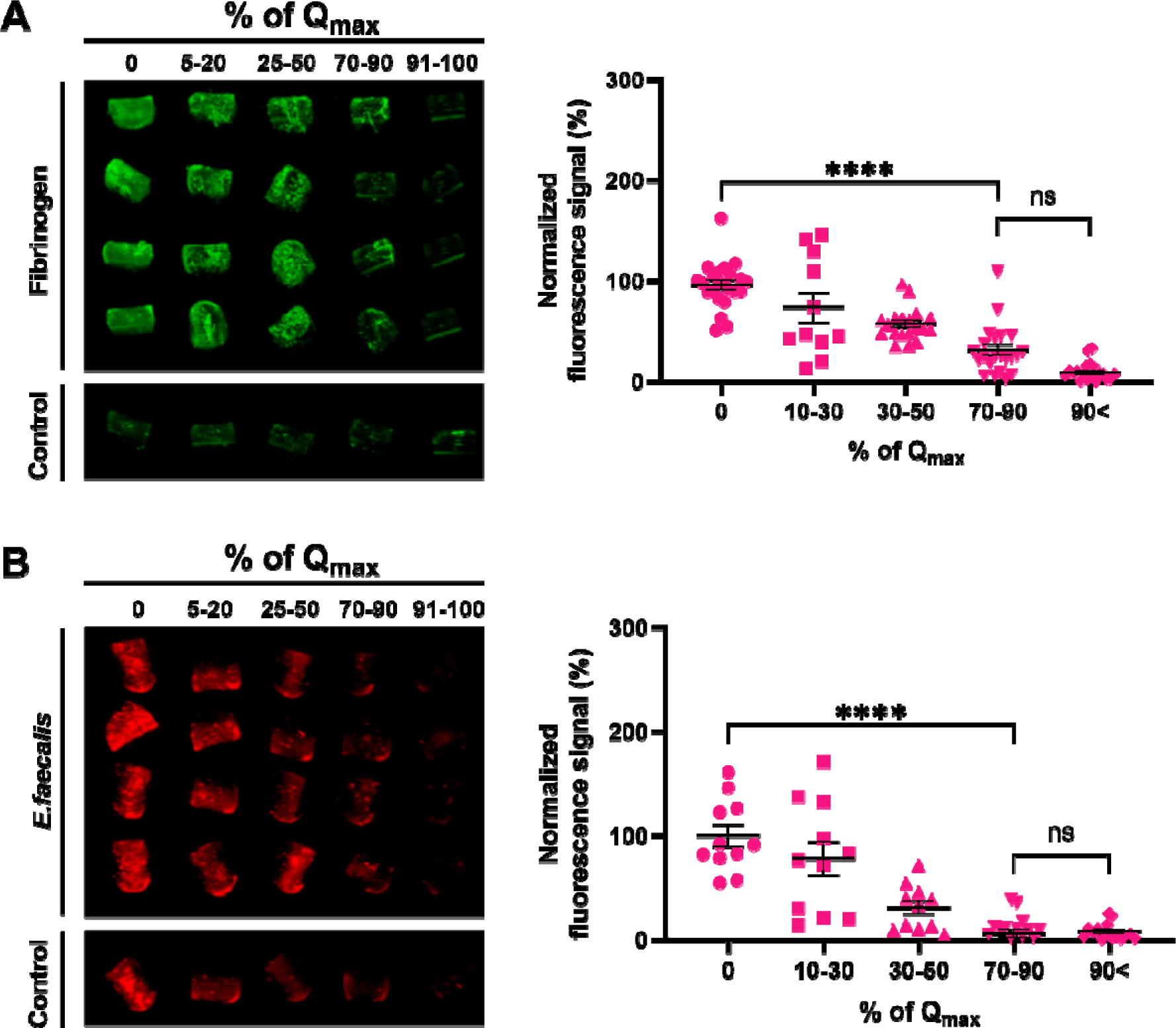
Immunolabelling of fibrinogen and *E. faecalis* on partially infused silicone catheter material. (A) Fibrinogen or (B) *E. faecalis* adhesion levels on silicone catheter material infused into various % values of Q_max_. The images on the left show the visualization of the samples while the plots on the right show the fluorescence quantification. At least three replicates with n = 4–5 each were conducted. In all graphs presented, the error bars represent the standard error of the mean. Statistical significance between groups was assessed using ANOVA. **** = P < 0.0001; and ns = not significant.

## 3. Discussion

Silicone material and silicone liquids are widely used in clinical settings, including as heart valves, breast implants, retinal tamponades, and syringe barrel lubricants, ^[40-44]^ due to their versatility and relative biocompatibility. Recent findings have shown that when free silicone liquid is infused into silicone catheter materials, there is a remarkable decrease in both fibrinogen and pathogen adhesion levels, both *in vivo* and *in vitro*. ^[13]^ This reduction in adhesion levels holds promise in effectively reducing the development of CAUTI without the need for antibiotics ^[13,45-47]^— a critical need given the rapid increase in resistant microorganisms.^[48-50]^ However, the potential for free silicone liquid to separate from the surface is a critical concern that must be addressed to increase the safety of these materials.

The potential for loss of the free liquid layer from liquid-infused system is well documented,.^[7-8,34,51]^ Under water, an infused surface can maintain a low sliding angle under both turbulent flow and laminar flow conditions for a little over an hour before experiencing a significant loss of the free liquid layer.^[52]^ However, it is well documented that when a surface with a free liquid layer is exposed to an air/water interface, the amount of liquid lost undergoes a substantial increase of 1–2 orders of magnitude, compared to being placed directly in water under flow.^[7]^ In addition, it is well understood that physical contact of a solid with a free liquid layer can also easily result in layer disruption and removal. ^[39]^ Given these facts, it is most likely that liquid-infused urinary catheters *in vivo* are being frequently subjected to conditions which can disrupt and/or remove any free liquid layer present on the catheter surface. Yet recent *in vivo* results demonstrate that infused catheters remain highly effective at resisting surface adhesion, despite the fact that they are unlikely to have a uniform liquid layer at the surface.

Current understanding of the mechanism of antifouling action of liquid-infused surfaces relies most heavily on the presence of a free and continuous liquid layer. It is thought that such a layer presents a physical barrier to contaminants and can deceive the mechanosensing mechanism of fouling organisms, preventing the initiation of their adhesive behavior.^[14-15,53-54]^ Recent RNA sequencing research has revealed that the introduction of silicone liquid to silicone solids leads to an upregulation of 10 distinct genes in *P. aeruginosa* while simultaneously downregulating a gene that may impede initial adhesion, although the precise mechanism behind this phenomenon remains unknown.^[37]^ Furthermore, the presence of the free liquid overlayer is thought to contribute to increased slipperiness on the infused surface, reducing the energy required for detaching fouling organisms and facilitating their release.^[8]^

More recently, understanding that liquid-infused silicone surfaces are more nuanced and dynamic than previously thought has been growing. Hatton *et al*. (2021) reported that when the free liquid layer was removed via washing with water, it would spontaneously regenerate over the following 360 hours (h), increasing linearly from ∼50 nm to ∼1µm. ^[16]^ Cai *et al*. (2021) demonstrated that free silicone liquid could spontaneously separate from the silicone solid at the edge of a water droplet.^[33]^ Wong *et al*. (2020) also showed that free molecules within the silicone solid could migrate to the surface in response to the presence of a water drop, but, importantly, also showed that they could return to the bulk after the droplet was removed.^[30]^ These results point to infused silicone surfaces as able to dynamically respond to changes in conditions. The results also suggest that liquid-infused silicone could perhaps be tuned to provide a desired response, such as fouling resistance, without an undesired response, such as surface liquid loss. Our findings provide further evidence of the dynamic and adaptive antifouling capacity of liquid-infused silicone materials. Even after removal of the bulk of the free liquid layer, we still found that these materials showed a significant reduction in fibrinogen and *E. faecalis* adhesion compared to non-infused controls. Importantly, removal of the free liquid layer resulted in a significant decrease in the amount of liquid that could be lost into the environment, suggesting that intentional removal of the free liquid layer may be an option to increase the safety of liquid-infused silicone catheters and other medical materials.

We further found that we could decrease the amount of liquid that could be removed from the surface by only partially infusing the catheter samples with silicone liquid. Due to the lower concentration of excess liquid in the system, the amount of free liquid available to accumulate at the surface is reduced.^[31]^ We found that at ∼80% of Q_max_, the amount of free silicone liquid that could be removed from the surface was significantly decreased compared to fully infused controls; and there was no statistically significant difference between fibrinogen and *E. faecalis* adhesion at this level. This is in agreement with previous work, which showed a significant reduction in *P. aeruginosa* adhesion even at infusion levels of 30% of their maximum. ^[16]^ Interestingly, Kolle *et al*. (2022) calculated that at a ∼20% loss of free liquid from a liquid-infused silicone bulk would result in a system in which a free liquid layer would no longer be able to re-form at the surface.^[39]^ Our results suggest that for medical applications, silicone infused to ∼80% of its Q_max_ value may contain just enough free liquid to create a dynamic interface that resists adhesion by proteins and microorganisms, while not enough to be easily lost into the environment.

Finally, although many investigations of liquid-infused silicones have examined their ability to repel bacterial adhesion,^[7-8,12,15-18,37,53-55]^ notably fewer have examined protein deposition.^[13-14,56]^ However, recent work has begun to reveal not only the critical role that proteins play in infection, ^[45-47]^ but also the ability of liquid-infused silicones to robustly resist their deposition.^[13]^ Here, we further show that protein deposition on liquid-infused surfaces can be modulated by adjusting the quantity of silicone liquid embedded in the polymer network. The ability to precisely modulate surface protein levels could potentially open new doors in a variety of fields, particularly materials engineering. In addition, the mechanism by which liquid-infused surfaces reduce protein deposition will be an important area of investigation going forward and may involve multiple interacting factors including masking of microscale defects and the neutralization of surface charges. ^[16,24,55,57-60]^

## 4. Conclusion

Improving the safety of silicone liquid-infused urinary catheters by reducing the amount of free silicone liquid available to be lost into host tissue is critical to their use in medical settings as alternatives to antibiotic coatings. Here, we removed the bulk of the free liquid layer from catheter samples by absorbing the liquid, resulting in a decrease in thickness of the liquid layer from ∼60 µm to <1 µm, and confirm that the result is a significant decrease in the amount of free silicone liquid that can be lost into the environment. Importantly, we find that removal of the liquid layer in this way does not prevent the surface from resisting deposition of fibrinogen and adhesion of *E. faecalis*, a host protein and uropathogen that have been shown to play a significant role in CAUTI. Our results suggest that minimal to no continuous free liquid layer may be required to be effective in medical applications such as device-associated infection prevention. Further investigation using catheter samples only partially infused with silicone liquid revealed that at ∼80% total infusion samples retained their ability to resist deposition and adhesion by fibrinogen and *E. faecalis,* respectively, while the amount of free liquid that could be removed from the surface was minimized. Both protein and bacterial adhesion were found to increase inversely with infusion levels below 80% suggesting that it may be possible to tune protein deposition and microbial adhesion using this method. Together, our results suggest potential benefits of incorporating the removal of the free liquid layer into the fabrication process of liquid-infused catheters as a method of both preserving antifouling properties while also improving safety.

## 5. Methods

### Infused silicone catheter material preparation

Silicone tubing (8060-0030, Nalgene^TM^ 50 Platinum-cured Silicone Tubing, Thermo Scientific, USA) was cut into 2-cm sections, before being fully submerged in 20 cSt silicone liquid (DMS-T12, Polydimethylsiloxane, Trimethylsiloxy terminated, 20cSt, Gelest, USA). 20cSt silicone liquid was chosen due to its use in recent *in vivo* CAUTI models.^[13]^ Any air bubbles in the tube lumen were removed to ensure uniform infusion. For LL and ØLL samples, the sections were removed from the silicone liquid after 5 days and excess liquid was allowed to flow out of the tube by holding it vertically for at least one minute or until all the excess liquid had dripped off the sample. These materials were then used as the LL samples; for the ØLL samples the sections were gently blotted on both the exterior and interior surfaces using absorbent cellulose wipe (Kimwipe, Kimberly-Clark Corp., USA) to absorb the free liquid. For partially infused samples, the sections were removed at defined points in the infusion process then weighed to determine % of Q_max_ value. All infused samples were allowed to equilibrate for at least 24 hours before further testing.

### Confocal imaging of infused samples with and without liquid layer

A mixture of 850 µg of BDP FL alkyne laser dye (D14B0, Lumiprobe, USA), 10 mL of dichloromethane, and 100g of silicone elastomer base (Dow SILGARD^TM^ 184 Clear, Dow, USA) were combined in a planetary centrifugal mixer (ARE-310, Thinky, USA) at 2000 rpm for 1 minute, followed by an additional mixing at 2200 rpm for 1 minute. The mixture was left in a desiccator overnight to remove any trapped gases. Next, 10 g of curing agent (Dow SILGARD^TM^ 184 Clear, Dow, USA; in a 10:1 ratio) was added to the resulting solution, which was mixed again in the centrifugal mixer using the same settings. Aliquots of 0.9 mL were then transferred into the square depressions (2.0 cm x 2.0 cm x 0.5 cm) of a mold master and were degassed for 2 hours and cured overnight at 70°C. To prepare the dyed silicone liquid for infusion, approximately 9 mg of pyrromethene (05971, Pyrromethene 597-8C9, Exciton, USA) was added to every 100 mL of silicone liquid and thoroughly mixed. The solution was filtered through a 0.45 µm filter to remove any particulates. The silicone squares were fully submerged in the infusion liquid for over 96 hours. Samples undergoing traditional infusion were placed vertically to drain off excess liquid. For samples requiring the overlayer to be stripped, they were placed vertically to allow excess infusion solution to drain off then gently dabbed on a Kimwipe (Kimwipe, Kimberly-Clark Corp., USA). Finally, the samples were imaged using a Leica Stellaris confocal microscope equipped with a white light laser, using an HC PL APO CS2 20x/0.75 mm objective lens. False coloring was added to the image with Clip Studio Paint (Clip Studio Paint, Japan).

### Sliding angle and droplet velocity tests

For the sliding angle tests, silicone tubing sections were placed on a tilt stage at 0°. A digital angle gauge (AccuMASTER 2 in 1 Magnetic Digital Level and Angle Finder, Calculated Industries) was affixed to the tilt stage to measure any changes in the angle. Using a pipette, a 20µl water droplet was deposited into the catheter’s lumen. The tilt stage was then gradually inclined until the droplet began to move off the tubing. The minimum angle at which the droplet began to move was then recorded. For the droplet velocity tests, the tilt stage was set at 30° and a digital camera (EOS Rebel T5 Digital SLR Camera; Canon; USA) was used to record the sliding of the droplet down the lumen. A frame-by-frame analysis was then used to determine the time the droplet took to travel from one end to the other.

### Free liquid removal of infused catheter samples

The dyed silicone liquid was prepared following the previously defined procedure. A standard curve of the dyed silicone liquid in MilliQ water (Millipore Milli-Q Direct 8 Water Purification System; 18.2MΩ-cm) was created by adding a fixed percentage of dyed silicone liquid in 10 ml of MilliQ water **(Figure S3)**. Afterward, 1 ml of toluene (108-883, Toluene anhydrous, Alfa Aesar, USA) was pipetted into the MilliQ water, and the mixture was manually shaken for 10 seconds and left to settle for at least a minute to separate into upper and bottom layers. The top layer was then carefully extracted and placed in a glass cuvette for spectrophotometer (840-277000, GENESYS™ 30 Visible Spectrophotometer, Thermo Fisher, USA) measurement. The absorbance of the samples was measured at 2nm intervals within the 350–650 nm range. A minimum of 20 samples per standard solution were used in the generation of the standard, adhering to the recommended number for establishing the limit of detection.^[61]^ The limit of detection, determined following the guidelines set forth by the Clinical and Laboratory Standards Institute, was calculated to be 0.0012%. The assessment of free liquid removal from catheter samples infused with dyed silicone liquid followed a consistent procedure. Instead of dispensing a fixed volume of liquid into MilliQ water, we immersed samples, prepared using the aforementioned infusion method, into 10 ml of MilliQ water. This immersion-withdrawal cycle was repeated ten times. Subsequently, the spectrophotometer readings were compared against a standard curve to quantify the amount of liquid extracted.

### Immunolabelling of fibrinogen and E. faecalis

Adhesion testing with human fibrinogen free from plasminogen and von Willebrand factor (Enzyme Research Laboratory #FB3) and *Enterobacter faecalis* (ATCC 47077) were conducted using previously described methods. ^[62]^ Briefly, after overnight incubation with fibrinogen in PBS (150µg/ml) or *E. faecalis* in human urine (supplemented with 20 mg/ml BSA), the catheters were fixed using 10% neutralized formalin, followed by blocking and staining steps. For fibrinogen, goat anti-Fg primary antibody (Sigma) was used in staining at a dilution of 1:1000, followed by Donkey anti-Goat IRD800 antibody (Invitrogen; 1:5000). For *E. faecalis,* rabbit primary antibody is used followed by Donkey anti-Rabbit IRD680 antibody (Invitrogen; 1:5000). After overnight drying at 4°C, the catheters were imaged using an Odyssey Imaging System (LI-COR Biosciences) to visualize and quantify the infrared signal.

### Statistical analysis

The statistical significance of the experimental results was evaluated using GraphPad Prism, version 7.03 (GraphPad Software, San Diego, CA). The data underwent an initial assessment to determine their adherence to Gaussian distribution. Based on the outcomes of these assessments, appropriate statistical tests were chosen. When the data exhibited Gaussian distribution, a one-way ANOVA was employed. Conversely, when the data did not conform to Gaussian distribution, the Kruskal-Wallis test was utilized. Significance levels on the graphs are denoted as follows: *p ≤ 0.05, **p ≤ 0.01, ***p ≤ 0.0001, and ****p ≤ 0.00001.

## Supporting Information

Supporting Information is available from the Wiley Online Library or from the author.

## Data Availability

All data produced in the present study are available upon reasonable request to the authors

## Acknowledgements

The authors thank Mr. Adrian Arias Palomo for technical editing; Dr. Emma Perry for her assistance with the confocal microscopy and Dr. Bill Halteman for statistical consulting. This work was supported by the National Institutes of Health grant under award number R01DK128805 and National Science Foundation grant no. CBET-1939710.

---

**Figure S1.**
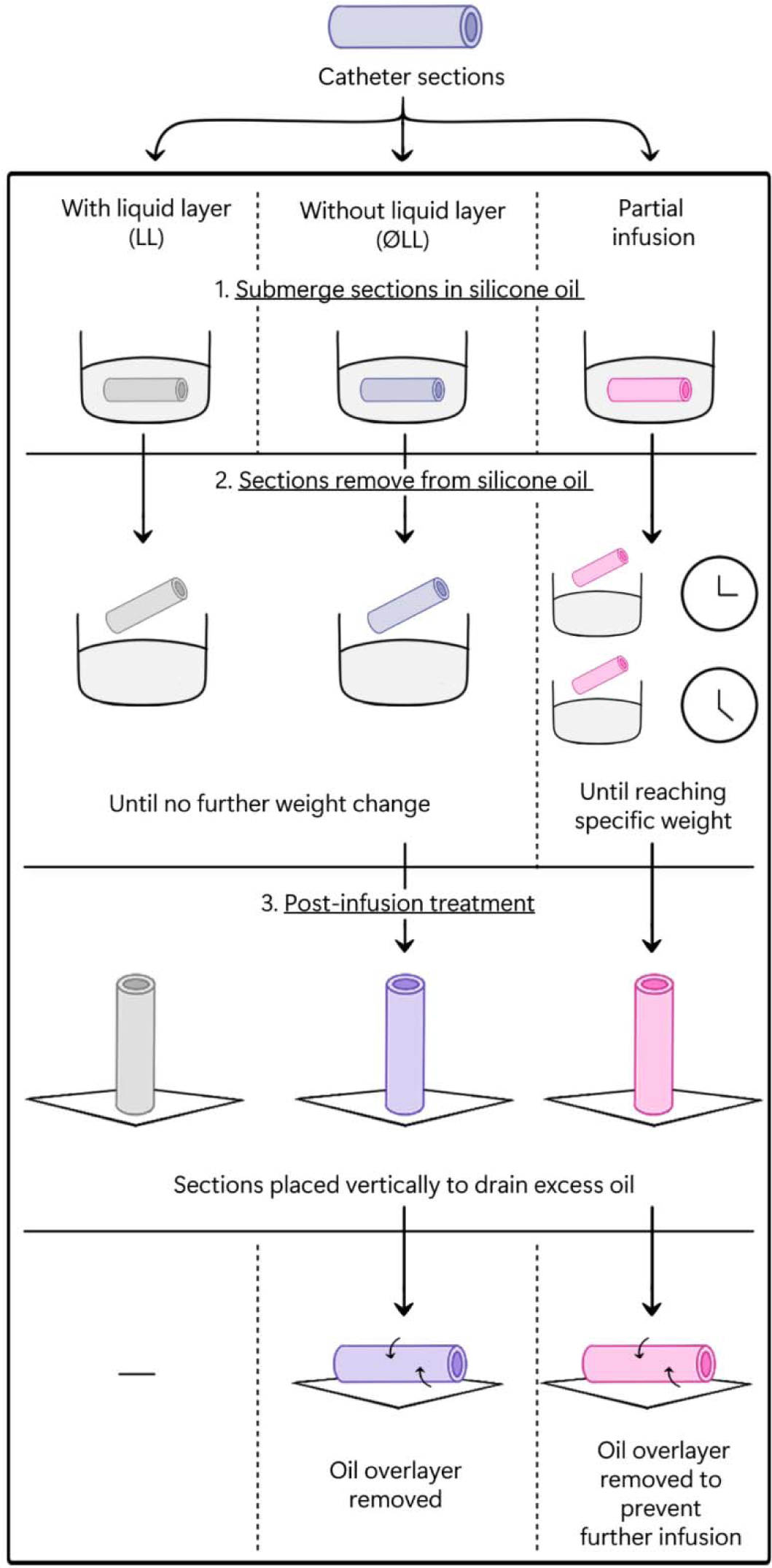
Infusion methods to fabricate infused catheters.

**Figure S2.**
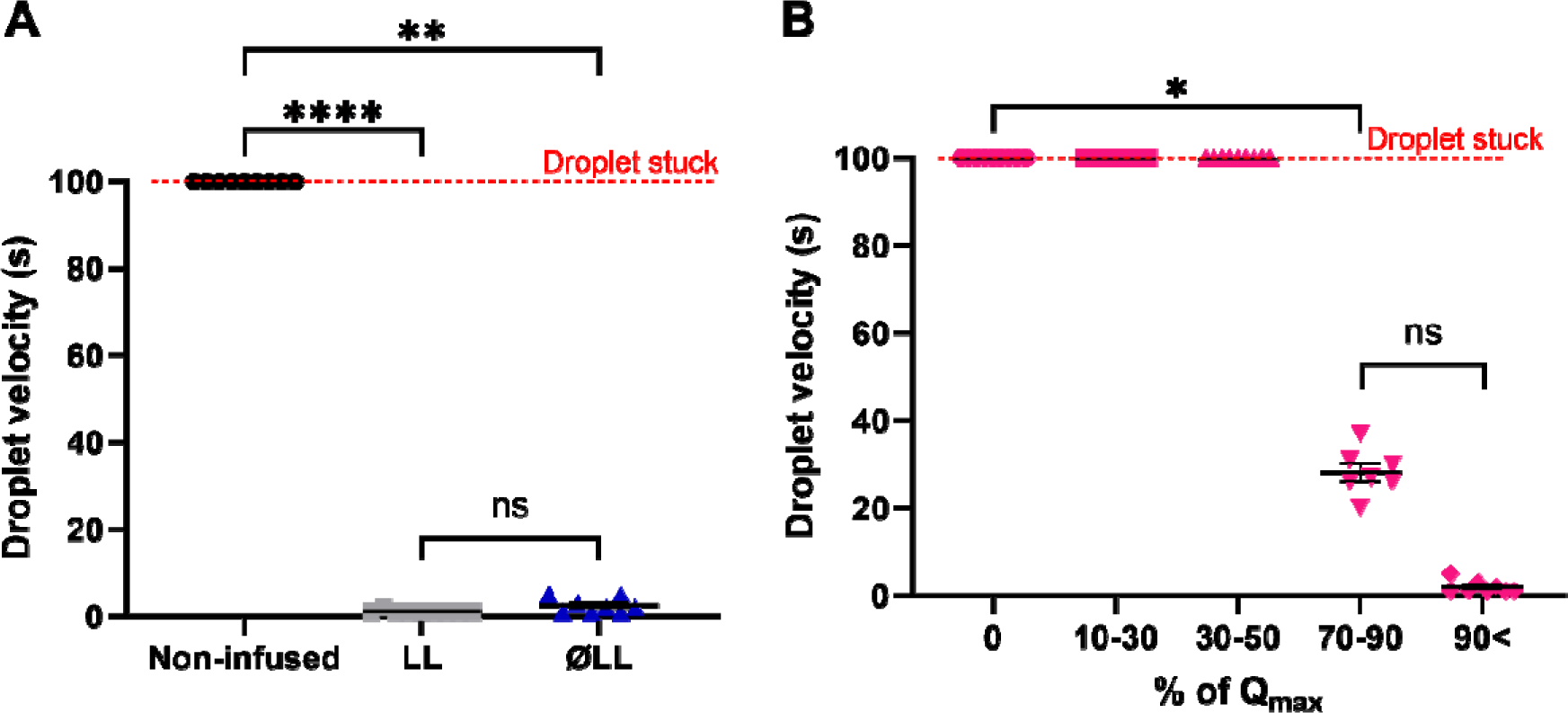
Droplet velocity on different infused catheter samples. Droplet velocity test results for **(A)** LL and ØLL silicone catheter samples; and **(B)** silicone catheter samples infused into various % of Qmax. In all graphs presented, the error bars represent the standard error of the mean (SEM). Statistical significance between groups was assessed using the Kruskal-Wallis test. **** = P < 0.0001; ** = P < 0.005; * = P< 0.05 and ns = not significant.

**Figure S3.**
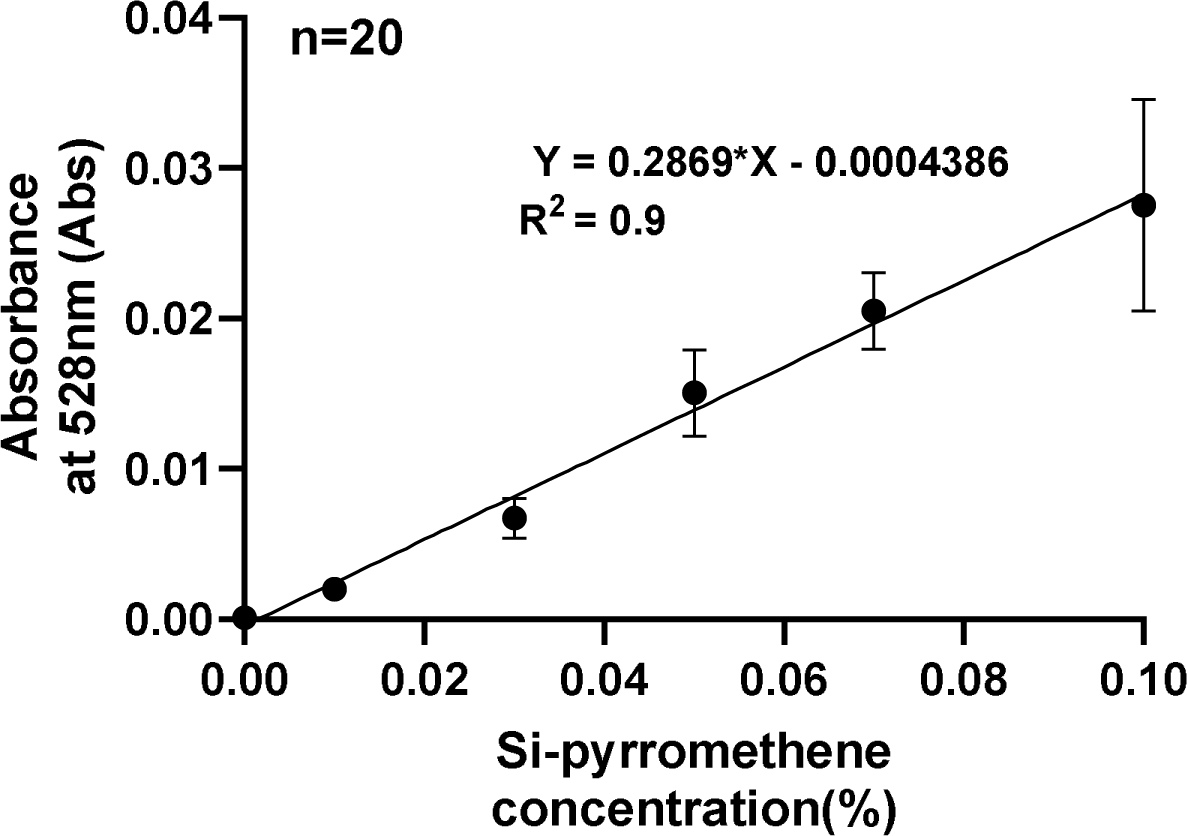
Standard curve for silicone liquid-pyrromethene mixture in toluene. Standard curve developed from silicone-pyrromethene mixture in toluene of known percentage. The error bars represent the standard error of the mean. n=20.

## Notes

### Competing Interest Statement

The authors have declared no competing interest.

