## Supplementary Information for "Removal of Free Liquid Layer from Liquid-Infused Catheters Reduces Silicone Loss into the Environment while Maintaining Adhesion Resistance"

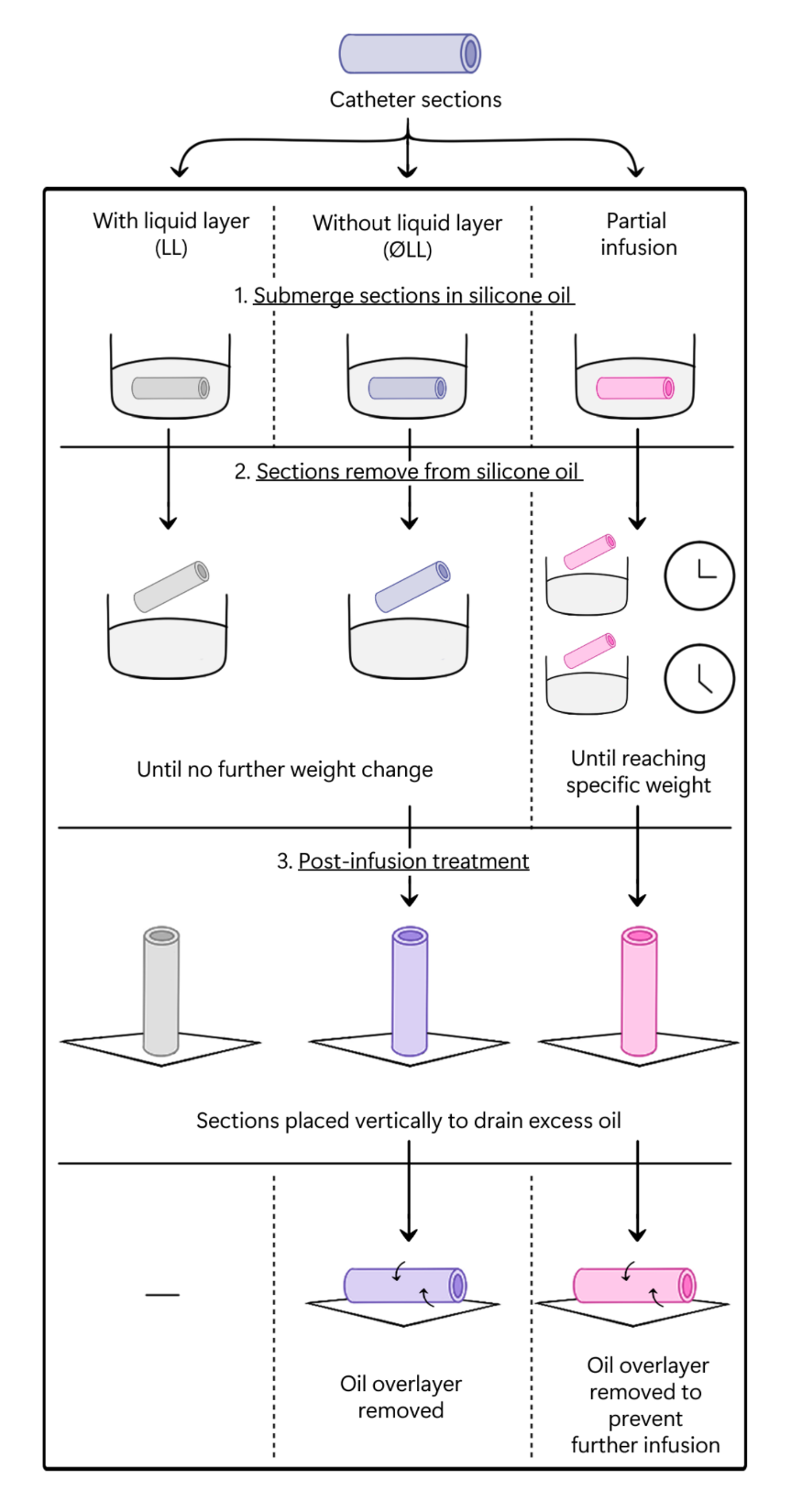

**Figure S1. Infusion methods to fabricate infused catheters.**

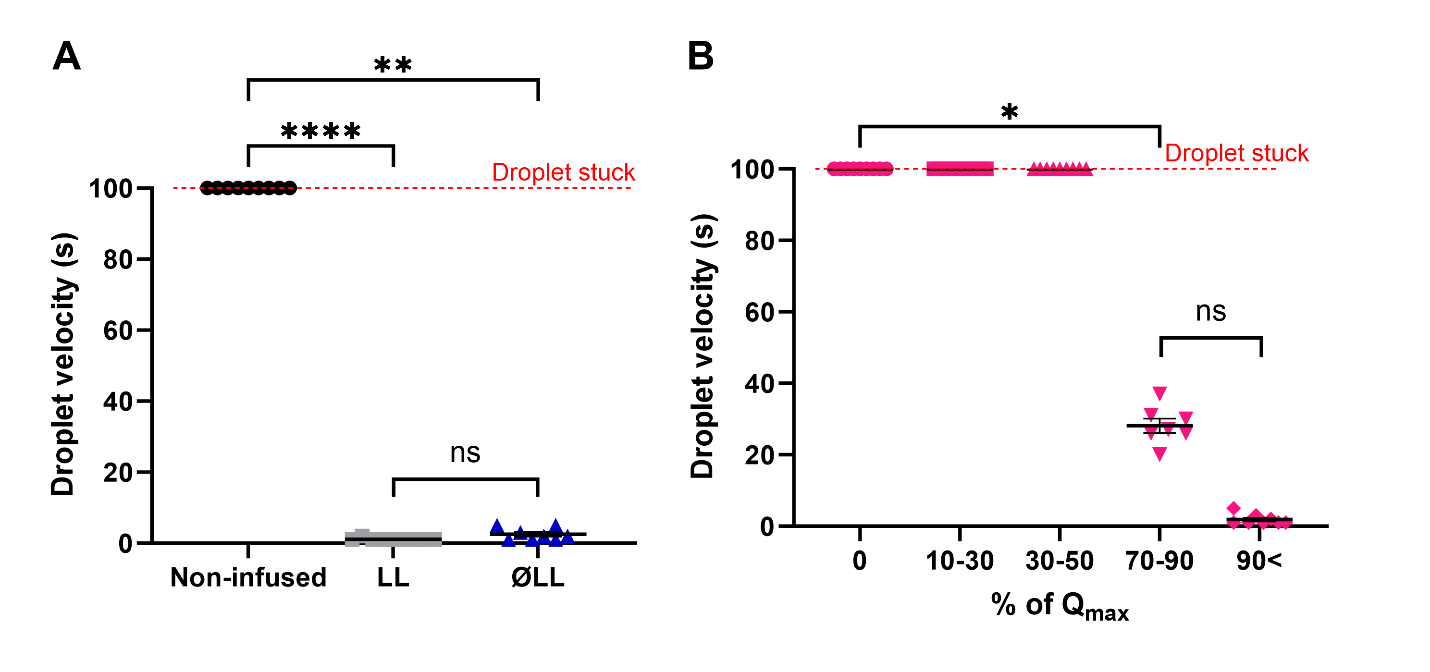

**Figure S2. Droplet velocity on different infused catheter samples.**  Droplet velocity test results for **(A)** LL and ØLL silicone catheter samples; and **(B)** silicone catheter samples infused into various % of Qmax. In all graphs presented, the error bars represent the standard error of the mean (SEM). Statistical significance between groups was assessed using the Kruskal-Wallis test. **** = P < 0.0001; ** = P < 0.005; * = P< 0.05 and ns = not significant.

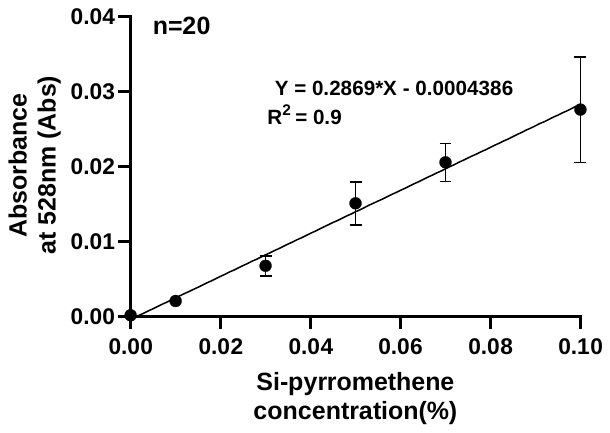

**Figure S3. Standard curve for silicone liquid-pyrromethene mixture in toluene.** Standard curve developed from silicone-pyrromethene mixture in toluene of known percentage. The error bars represent the standard error of the mean. n=20.
